# Development and Validation of Two In-house, Low-Cost SARS-CoV-2 Detection Assays

**DOI:** 10.1101/2020.05.18.20105510

**Authors:** Fatimah S. Alhamlan, Ahmed A. Alqahtani, Dana M. Bakheet, Marie F. Bohol, Sahar I. Althawadi, Maysoon S. Mutabagani, Reem S. Almaghrabi, Dalia A. Obied

**Affiliations:** Department of Infection and Immunity, King Faisal Specialist Hospital and Research Center, Riyadh, Saudi Arabia; College of Medicine, Alfaisal University, Riyadh, Saudi Arabia; Department of Genetics, King Faisal Specialist Hospital and Research Center, Riyadh, Saudi Arabia; Department of Pathology and Laboratory Medicine, King Faisal Specialist Hospital and Research Center, Riyadh, Saudi Arabia; Department of Medicine, King Faisal Specialist Hospital and Research Center, Riyadh, Saudi Arabia

**Author notes:** **Corresponding author**. Fatimah S. Alhamlan.

**Keywords:** SARS-CoV-2, COVID19, Diagnostic tests, rtPCR, PCR

## Abstract

**Background:** One major challenge for detecting the virus that causes COVID-19 is commercial SARS-CoV-2 testing kit or reagent availability. To allow every laboratory or hospital access to an in-house assay, we developed two low-cost SARS-CoV-2 detection assay protocols using in-house primers and reagents/equipment on hand in most biology or diagnostic laboratories: a SYBR Green–based RT-PCR and PCR assays. RNA extraction has also become a major bottleneck due to limited supplies and the required labor. Thus, we validated alternative RNA extraction protocols.

**Methods:** SARS-CoV-2 genome sequences deposited into the GISAID database were retrieved to design and synthesize in-house primers. Forty patient samples were collected by nasopharyngeal swab, coded, and used to develop and validate the assay protocols. Both assays used TRIzol and heat-processing techniques to extract RNA from patient samples and to inactivate the virus; thus, testing was conducted in a conventional biosafety level 2 laboratory.

**Results:** The sensitivity and specificity of the primers were evaluated using samples previously confirmed positive for SARS-CoV-2. The positive amplicons were sequenced to confirm the results. The assay protocols were developed, and the specificity of each PCR product was confirmed using melting curve analyses. The most accurate heat-processing technique for primers with short amplicon lengths was 95 °C for 15 mins. Of 40 samples, both the SYBR Green–based quantitative RT-PCR assay and the PCR assay detected SARS-CoV-2 target genes in 28 samples, with no false-positive or false-negative results. These findings were 100% concordant with those of the diagnostic laboratory that tested the same samples using a Rotor-Gene PCR cycler with an Altona Diagnostics SARS-CoV-2 kit (R2=0.889).

**Conclusions:** These approaches are reliable, repeatable, specific, sensitive, simple, and low-cost tools for the detection of SARS-CoV-2 in a conventional biosafety level 2 laboratory, offering alternative approaches when commercial kits are unavailable or cost ineffective.

## 1. Introduction

In March 2020, the World Health Organization declared a global emergency for a pandemic caused by the novel coronavirus SARS-CoV-2. This virus belongs to the *Coronaviridae* family, which commonly infects humans and mammals, and is an enveloped, non-segmented, positive-sense RNA. This family of viruses has caused two previous epidemics: severe acute respiratory syndrome coronavirus and Middle East respiratory syndrome coronavirus infections [1, 2]. Although these viruses belong to the same family, they rely on different mechanisms and have different manifestations in the human host. The newly emerged virus was first detected in Wuhan, China, and as of April 4, 2020, it had infected more than 1 million people worldwide and caused the deaths of 58,937 individuals [3].

To slow the spread of the virus and to contain the infection, countries across the globe are attempting to screen millions of individuals, including all health care personnel and their contacts, patients who have symptoms associated with infection by SARS-CoV-2 and their contacts, individuals who have come into contact with an infected patient, and asymptomatic individuals. People who carry the virus without showing any symptoms pose a major silent threat to public health because they are unknowingly spreading the virus. However, not all countries, hospitals, or laboratories have access to testing kits or to the reagents and supplies that are currently in high demand with which they could make their own kits. In addition, many hospitals and diagnostic laboratories are struggling with an overwhelming number of samples that have been collected but still require testing. Therefore, the aim of the present study was to develop and validate two inexpensive assays to detect SARS-CoV-2 in patient samples in almost any laboratory or hospital in a timely manner and without commercial, ready-made extraction or detection kits. Both assays can use a TRI reagent-based RNA extraction or heat-processed samples for RNA extraction. The RNA extraction procedure is followed by cDNA synthesis then a SYBR Green–based RT-PCR or conventional PCR assay that requires only basic science techniques and equipment. Here we provide two validated protocols for safely detecting SARS-CoV-2 in a conventional biosafety level 2 laboratories within a limited budget by using these easily performed assays.

## 2. Materials and Methods

### 2.1 Sample collection and ethical considerations

This study was performed in compliance with all applicable national and international ethical guidelines for conducting research on human participants, including in accordance with The Code of Ethics of the World Medical Association (Declaration of Helsinki), and was approved by the institutional review board at King Faisal Specialist Hospital and Research Centre IRB # 2200021. This board also granted a waiver for obtaining informed consent owing to the use of deidentified samples for this study.

Samples were collected from each patient using a nasopharyngeal swab and then coded by the Microbiology Section of the Pathology and Laboratory Medicine Department at King Faisal Specialist Hospital and Research Centre (KFSHRC). Of these deidentified samples, those from 40 patients who were tested for SARS-CoV-2 infection were included in the present study.

### 2.2 RNA extraction

Three alternative protocols for RNA extraction were validated to accommodate the needs and available resources of various laboratories.

#### 2.2.1 TRI Reagent-based RNA extraction (manual)

To inactivate the virus, 300 μL from the patient sample was added to 700 μL of TRI reagent (Sigma-Aldrich, USA) and mixed in a biosafety cabinet in a negative pressure room. The tube was inverted 10 times and kept at room temperature for 5 min. Because this step inactivates the virus, it was safe to continue the extraction process in a biosafety level 2 laboratory. Next, 200 μL of chloroform was added, and the tube was inverted five times and then incubated for 3 min at room temperature. The sample was centrifuged at 12,000 × *g* for 10 min at 4 °C. The aqueous layer, which contains RNA (approximately 500 μL), was transferred to a new 1.5-mL tube, and 500 μL of isopropanol was added. The tube was inverted five times and then centrifuged at 12,000 × *g* for 10 min at 4 °C. The supernatant was removed, and 1 mL of 75% ethanol was added. The sample was centrifuged at 7500 × *g* for 5 min at 4 °C. The supernatant was removed, 1 mL of 75% ethanol was added to wash the sample, and was again centrifuged at 7500 × *g* for 5 min at 4 °C. The supernatant was discarded and the pellet was air-dried. Next, 15 μL of AVE (elution) buffer was added to re-suspend the RNA. However, RNase-free water can be used instead of AVE buffer. We then added 1 μL of RNaseOut (Invitrogen, USA) to inhibit RNases and to protect the RNA from degradation. The RNA concentration and purity were determined with a NanoDrop spectrophotometer (Thermo Fisher Scientific, USA).

#### 2.2.2 TRI Reagent-based RNA extraction (automated protocol)

To establish a high-throughput RNA extraction technique for mass testing, we modified the TRI reagent-based extraction for use on a Hamilton Microlab STAR Liquid Handling System. Magnet beads (ChargeSwitch^;^ Invitrogen) were used for purifying the total RNA from the TRI reagent lysis step. To obtain high-quality total RNA, 300 μL from the patient sample was added to 700 μL of TRI reagent (Sigma-Aldrich, USA) and mixed in biosafety cabinet in a negative pressure room using a 96-well plate. The plate was placed on a shaker for 1 min to homogenize the solution. Next, 200 μL of chloroform was added, and the plate was placed on a shaker for 1 min to again homogenize the solution and then incubated for 3 min at room temperature. The plate was centrifuged at 12,000 × g for 10 min at 4 °C. The automation process started at this step, in which the plate was placed in the Microlab STAR Liquid Handling System. Because obtaining the aqueous layer, which contains RNA (approximately 500 μL), is the most critical step in the extraction process, using an automated and robust robot improves accuracy by eliminating errors that can occur during manual processing and can extract pure RNA from a 96-well plate in less than 15 min.

#### 2.2.3 Extraction-free protocol (heat-processed samples)

From the nasopharyngeal swab (stored in Viral Transport Medium), 300 μL of the sample was placed in a separate tube. Samples were heated at 95 °C for 15 min. The reaction tubes were placed on ice to cool before proceeding. Samples were centrifuged at 7500 × *g* for 5 min to remove any debris. The supernatant was then transferred using a pipette into a new tube to be used for cDNA synthesis and the subsequent PCR reactions. The RNA concentration and purity were determined with a NanoDrop spectrophotometer (Thermo Fisher Scientific).

### 2.3 cDNA Synthesis

The synthesis of cDNA is required to prepare patient samples for subsequent PCR reactions. Total RNA extract (8 μL of approximately 50 ng/μL) was added to 1 μL of random hexamer primers and 1 μl of 10 mM dNTP Mix (Thermo Fisher Scientific). The mixture was incubated at 65 °C for 5 min and then placed on ice. Next, a master mix was prepared that consisted of 2 μL of 10× Reverse Transcriptase (RT) buffer, 4 μL of 25 mM MgCl_2_ μL of 0.1 M dichlorodiphenyltrichloroethane (DDT), 1 μL of RNaseOut, and 1 μL of the reverse transcriptase enzyme (SuperScriptIII RT; 200 U/μL; Thermo Fisher Scientific). The master mix (10 μL) was added to the RNA extraction mix (10 μL) and mixed by manually tapping and pulse-spinning. Samples were incubated at room temperature (approximately 25 °C) for 10 mins and then incubated at 50 °C for 50 min. The reaction was terminated at 85 °C for 5 min and then placed on ice. After pulse-spinning the sample, 1 μL of RNase H (to remove the RNA template from RNA-cDNA hybrid) was added, and the sample was incubated at 37 °C for 20 min. The quality and the quantity of the cDNA samples were determined using a NanoDrop spectrophotometer.

### 2.4 Nucleic acid purity assessment

The quality and the quantity of the RNA and cDNA samples were also determined using a NanoDrop spectrophotometer. The ratio of sample absorbance at wavelengths of 260 and 280 nm were obtained to assess the nucleic acid purity (approximately 2.0 for RNA and approximately 1.8 for cDNA).

### 2.5 Designing primer sets specific for SARS-CoV-2

More than 150 complete SARS-CoV-2 genome sequences were retrieved from the GISAID database (https://www.epicov.org/epi3/cfrontend#lightbox-814829872) and were aligned using the Clustal W algorithm of the MegAlign module to identify the conserved regions using DNAStar software (DNASTAR; Madison, WI). Genes for the major structural proteins of SARS-CoV-2, such as S, E, M, ORF1a and N, were targeted. Oligonucleotide primers were designed for the SARS-CoV-2 S genes to ensure maximal efficiency and sensitivity. The desired primers were designed using the consensus sequences from different SARS-CoV-2 isolates from around the world, and the Wuhan first genome sequence was used as a reference (Accession No. MN908947).

Because a successful PCR assay requires efficient and specific amplification, the primers were assessed for several properties, including melting temperature, secondary structure, and complementarity. Primers need to have a GC content of 50%-60%, a melting temperature between 50 °C and 65 °C, 50 mM salt concentration, and 300 nM oligonucleotide concentrations. The specificity of the primers and final sequences were verified using in silico prediction analyses with the online Basic Local Alignment Search Tool (BLAST) (https://blast.ncbi.nlm.nih.gov/Blast.cgi). None of our designed primers showed genomic cross-reactivity with other viruses, the human genome, or other probable interfering genomes in the BLAST database analysis. Primers were synthesized in-house at the Oligonucleotide Synthesis Unit of the Genetics Department at KFSHRC.

### 2.6 SYBR Green RT-PCR assay

The ΔΔCt method of relative quantification using RT-PCR with SYBR Green detection was used for this study. To detect SARS-CoV-2 target genes, comparative quantitative RT-PCR was performed using SYBR Green fluorescent dye, which binds double-stranded DNA by intercalating between the DNA bases. The 7500 Fast Real-Time PCR system and software (Applied Biosystems, California, USA) were used.

The target SARS-CoV-2 genes included N, S, E, and RdRp. In addition, RNase P, which is a housekeeping gene, was used as an internal positive control. Primers targeting the N, S, E, RdRp and RNase P genes were designed and synthesized in-house by the Oligonucleotide Unit of KFSHRC (Table 1). SYBR Green RT-PCR assays using these primers generated in-house were conducted for all patient samples that were included in this study. To amplify cDNA in the patient samples, a master mix was prepared with 12.5 μL of SYBR Green master mix, 0.5 μL of forward primer (20 μM), 0.5 μL of reverse primers (20 μM), 2 μL of cDNA, and 4.5 μL of water (for a total of 20 μL). The amplification conditions were 50 °C for 20 secs and 95 °C for 10 mins followed by 45 cycles of 95 °C for 3 s with 55 °C for 30 s. At the end of each reaction, the cycle threshold (Ct) was acquired at the level that reflected the best kinetic PCR parameters.

**Table 1.**
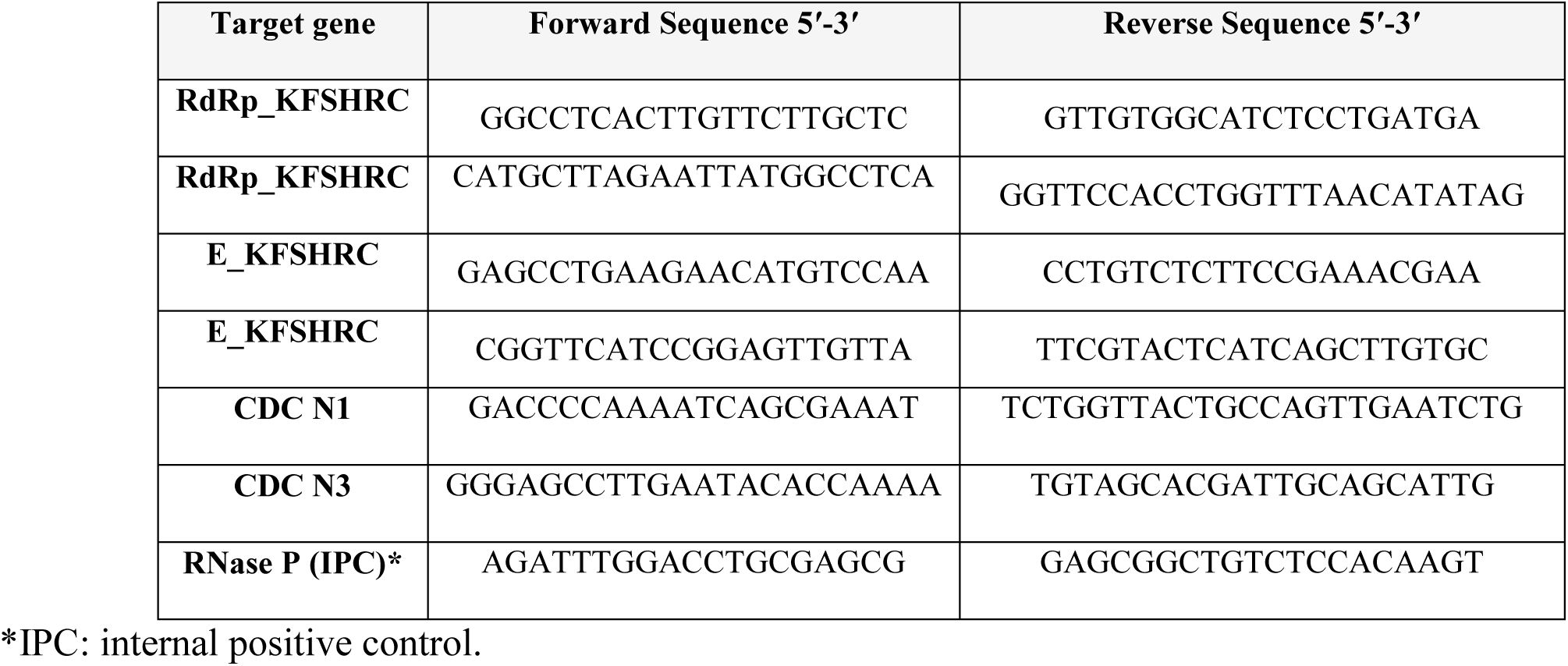
Real-Time-PCR primer sets for SARS-CoV-2 detection using the SYBR Green-based assay

#### 2.6.1 Melting curve analysis

SYBR Green is released when a PCR product is denatured, which results in a rapid increase in absorbance intensity (fluorescence signal) followed by a decrease in the signal. When an amplified PCR product is specific, a melting curve generated using that product should provide a single peak that corresponds to that PCR product. Thus, to check the specificity of each PCR product, we conducted a melting curve analysis at the end of each PCR assay. Each product sample was analyzed once. For the melting curve, the fluorescence signal of each PCR product was monitored continuously as the temperature was increased to 95 °C for 15 s, decreased to 60 °C for 1 min and then increased again to 95 °C for 30 s and 60 °C for 15 s.

### 2.7 PCR

The PCR primer sets for SARS-CoV-2 detection were designed and synthesized at KFSHRC. The primer sets used are listed in Table 2. For a total reaction volume of 30 μL was prepared that included 3 μL of the SARS-CoV-2 cDNA, first round primers (0.5 μL each/20 μM), 15 μL of GoTaq Green Master Mix (Promega; Madison, WI, USA), and 11 μL of DNase/RNase-free water. The PCR cycle was performed as follows: 95 °C for 5 min; then 40 cycles of 95 °C for 30 s, 58° C for 30 s, and 72 °C for 45 s; followed by 72 °C for 5 min. Controls (positive and negative) were used. The products from the PCR were separated using 2% agarose gel electrophoresis and visualized using ultraviolet light (Gel Doc EZ Gel; Bio-Rad). The remaining PCR products were purified using AMpure XP and then sent for Sanger sequencing to the Core Facility of the Genetics Department at KFSHRC. The analyzed sequences matched the WH-human1 SARS-CoV-2 sequence (MN908947).

**Table 2.**
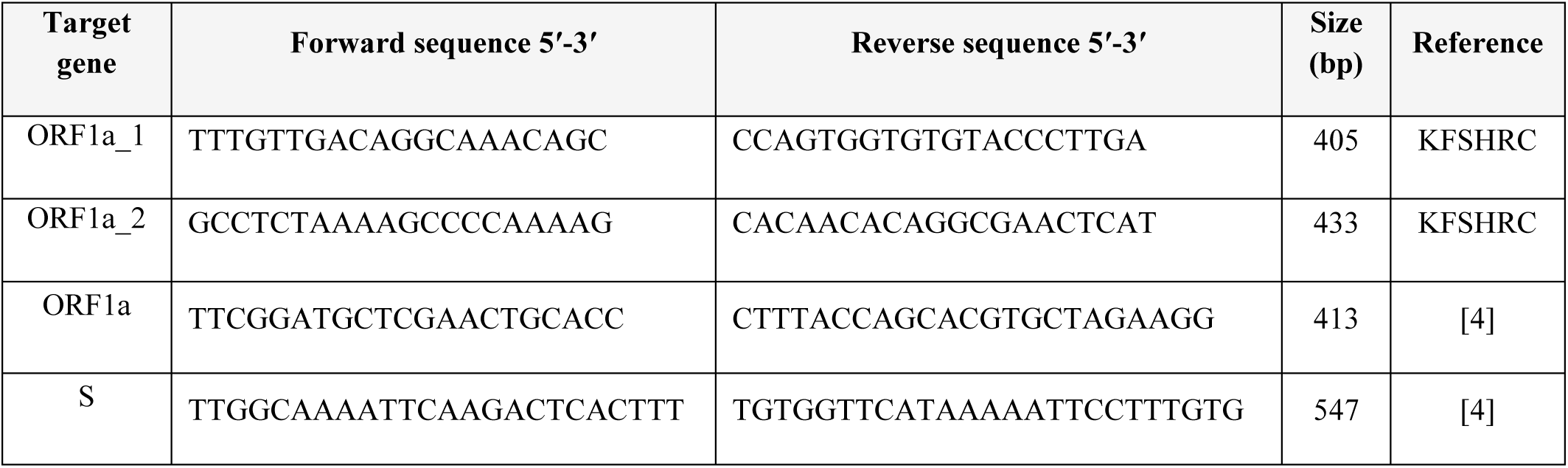
PCR primer sets for SARS-CoV-2 detection

## 3. Results

### 3.1 Total RNA extraction using three different approaches

During RNA extraction using TRI reagent, we obtained an average of 20 ng/μl from each sample. Using the heat-processing technique, the RNA concentration was higher but the purity was lower. Most importantly, none of the three extraction methods negatively affected the PCR results.

To evaluate the RNA extraction method, three RNA samples extracted with TRI reagent at the research laboratory were sent to a diagnostic laboratory for testing. The results showed that the Ct values for the RNA extracted using these methods were similar and also indicated the compatibility of the RNA extraction procedure using TRI reagent with the Rotor-Gene system using the Altona Diagnostics kit.

### 3.2 Detection of SARS-CoV-2 in test samples using the SYBR Green assay

For a valid diagnostic test, the sample was considered positive for SARS-CoV-2 when the cycle threshold growth curves for both target genes crossed the threshold line within 40 cycles (<40.00 Ct) regardless of whether the RNase P growth curve also crossed the threshold within 40 cycles. A sample was considered negative for SARS-CoV-2 when the cycle threshold growth curves for both targets did not cross the threshold line within 40 cycles (>40.00 Ct) but the RNase P growth curve did cross the threshold line within 40 cycles. Figure 1 shows a representative RT-PCR amplification plot. The results of our developed SYBR Green assay protocol indicated that of the 40 nasopharyngeal samples tested, SARS-CoV-2 was detected in 28. This result was 100% concordant with that of the diagnostic laboratory at KFSHRC that had originally tested the samples. Figure 1 depicted a representative amplification plot the showed two different target genes, internal positive control and negative control. Figure 2 showed the distribution of Cycle Threshold (Ct) Values by RT PCR assays. Altona Diagnostic PCR assay detected SARS-CoV2 mostly in earlier cycles than with SYBR-Green RT-PCR, though there was no significant difference.

**Figure 1.**
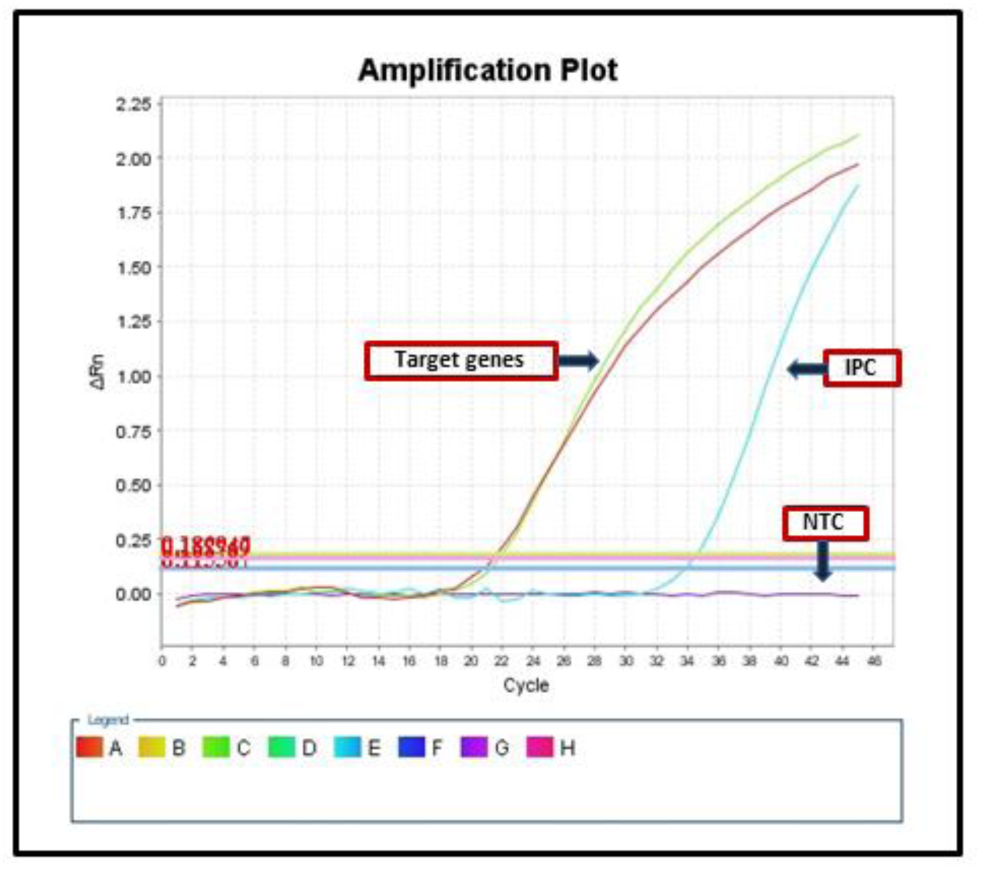
Representative Amplification Plot for Real-time PCR Using SYBR Green and the ΔΔCt Method of Relative Quantification. Plot showing the Ct value of two target genes, an internal positive control (IPC) and a no-template control (NTC). The lower the Ct value, the greater the sensitivity. ΔRn indicates Rn (the reporter signal normalized to the fluorescence signal of Applied Biosystems ROX Dye) minus the baseline; ΔRn is plotted against PCR cycle number.

**Figure 2.**
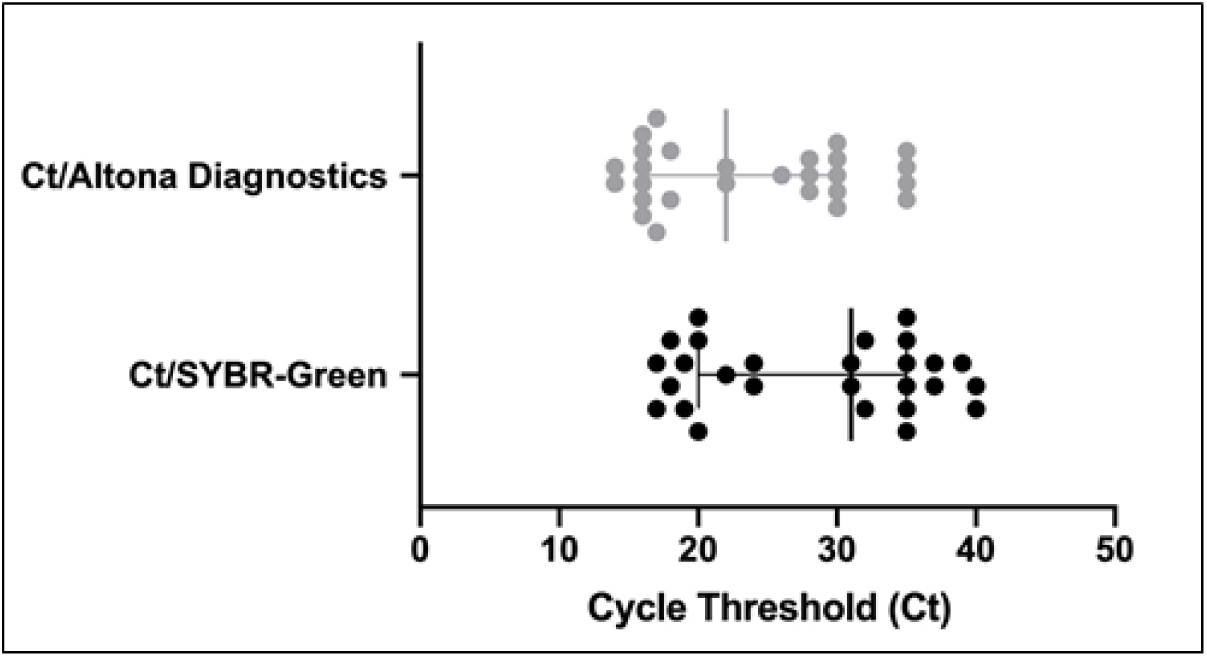
Distribution of Cycle Threshold (Ct) Values by RT PCR assays. Altona Diagnostic PCR assay detected SARS-CoV2 in earlier cycles (Mean=24) in comparison to SYBR-Green RT-PCR (Mean=28) however there was no significant differences.

**Figure 2.**
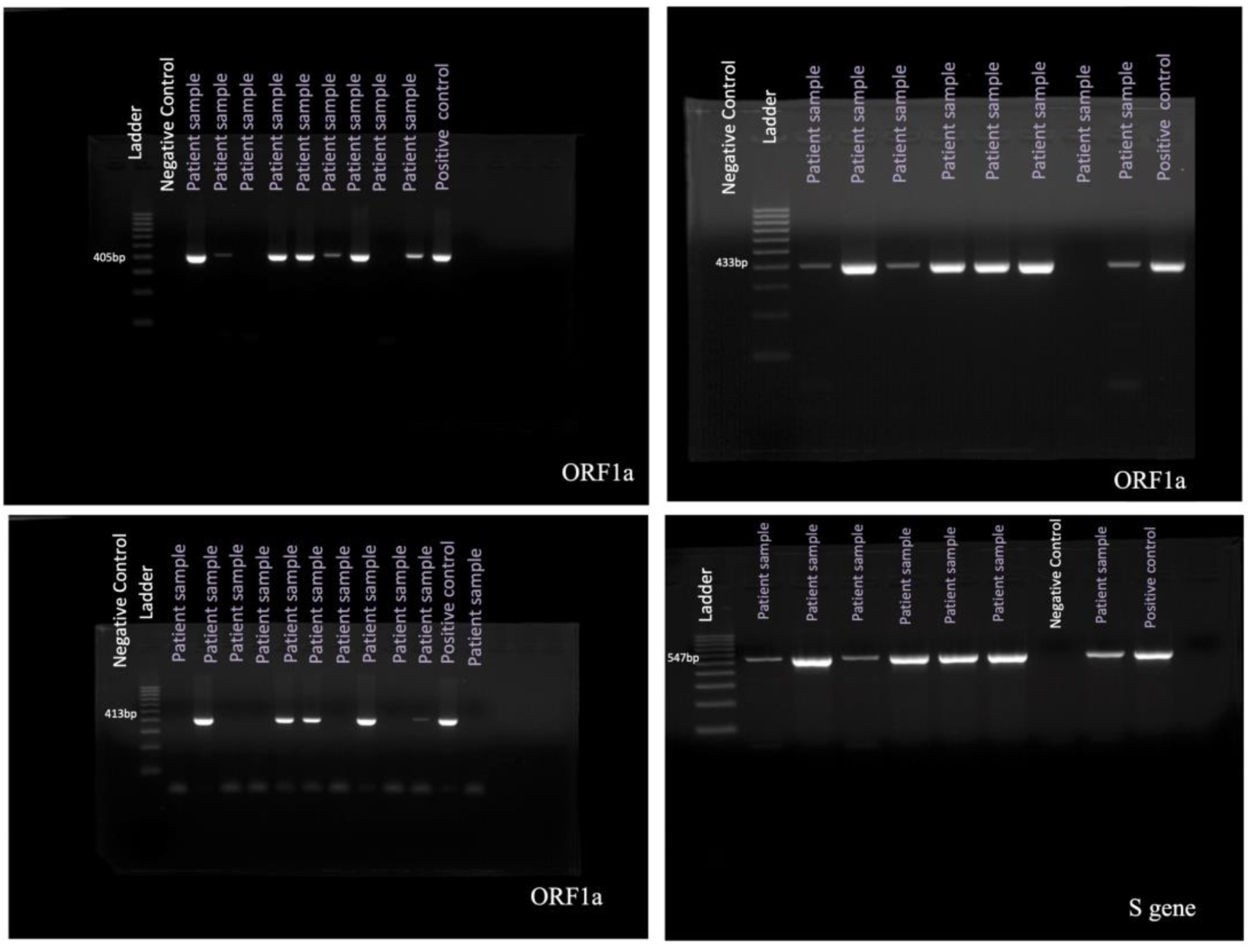
**Representative images of** PCR Detection of the SARS-CoV-2 ORF1a and S Genes. Reaction products tested on an agarose gel stained with ethidium bromide to confirm the results. DNA ladder is 100 base pairs (bp).

### 3.3 Detection of SARS-CoV-2 in test samples using PCR

For a valid diagnostic test using PCR, the following conditions must be met. For positive control samples, the bands with the corresponding amplicon size must be visualized, whereas for negative samples, the band must not be present. In addition, the positive control must be positive; if this test failed, the experiment was repeated. The test for the negative control (DNase/RNase-free water) must be negative; if this test failed, the experiment was repeated.

Consistent with the results of the SYBR Green assay, of the 40 tested nasopharyngeal samples, SARS-CoV-2 was detected in 27 samples. Figure 2 shows representative results of the PCR assay.

### 3.4 Test accuracy and validation

Parallel tests to detect SARS-CoV-2 were conducted by a diagnostic laboratory accredited by the College of American Pathologists and our research laboratory to validate the accuracy of the developed protocol. Both labs ran 28 identical deidentified nasopharyngeal samples. The research laboratory used SYBR Green and PCR assay protocols, whereas the diagnostic Molecular laboratory at the Department of Pathology have utilize either EZ1 Virus Mini Kit v2.0 (QIAGEN) or Abbott *m*2000 SP System with either DNA or RNA virus kit for extraction and the Rotor-Gene RT-PCR cycler (QIAGEN) with a commercial kit from Altona Diagnostics.

The results of the laboratories were 100% concordant. No false-positive or false-negative test results were reported using the two in-house assay protocols.

To determine the limit of detection, 10-fold serial dilutions using a plasmid with a known copy number (104/μL) were tested. Real time PCR and PCR were used to assess the sensitivity of the assay. The limit of detection is the lowest concentration of the diluted sample that returns a positive result. Our results showed that the limit of detection for our assays was 10 copies/μL.

The results from both laboratories were presented as either positive [Ct <40] or negative [Ct >40] for the detection of SARS-CoV-2. The Ct values in the positive samples ranged from 16 to 38. All PCR products were examined using electrophoresis of an agarose gel stained with ethidium bromide in standard TBE buffer. All of the detected positive samples showed a well-defined specific band with results consistent between the laboratories (i.e., faint bands for late Ct values). In addition, the melting curve analyses showed a single peak for each positive sample.

## 4. Discussion

Although commercial viral RNA extraction kits are available and are widely used in most laboratories, the aim of the present study was to use basic biology techniques that can be adopted by hospitals or laboratories worldwide, especially given that the availability of commercial kits or specialized reagents may be at risk. Thus, we developed and validated two low-cost, easily performed assays for detecting SARS-CoV-2: (1) a SYBR Green RT-PCR assay and (2) PCR assay. The tests can be conducted using equipment and reagents already found in most pathology or diagnostic laboratories.

The heat-processed RNA extraction procedure showed promising results. This finding is in agreement with a recent study that used this approach with various temperatures and times (5 min at 95°C; 10 min at 95°C; 5 min at 98°C; 10 min at 98°C) [5]. The present study found that the optimum temperature for robust results was 95 °C for 15 min. It is important to note that short amplicon sequences are critical for successful detection using heat-processed samples because it is the heat that shears the RNA.

Testing with RT-PCR has been considered the gold standard for diagnosing COVID-19, with infected individuals identified by the successful amplification of the viral genome obtained using nasopharyngeal swabs. However, recent studies have stated some concerns regarding the specificity of the RT-PCR tests for patients with COVID-19 [6]. Indeed, inaccurate results, especially a false-negative, have dire implications for subsequent community transmission of the virus [7]. Therefore, the inclusion of two SARS-CoV-2 genes for accurate detection has been advised [8]. In the present study, we designed sets of primers that target the E and RdRp genes of the virus.

SYBR Green quantitative PCR is widely used because of the ease in assay design and its relatively low setup and running costs. Unlike TaqMan fluorescent probes, SYBR Green dye intercalates into double-stranded DNA to monitor the amplification of the target gene specifically initiated by gene-specific primers [9]. However, one drawback of the SYBR Green assay is that the dye is nonspecific, and this lack of specificity can generate false-positive signals if nonspecific products or primer-dimers are present in the sample. However, including a melting curve analysis at the end of each PCR assay to determine the specificity and efficiency of each RT-PCR reaction will ensure the accuracy of the results when multiple peaks or primer-dimers are not observed. Another potential drawback of the SYBR Green assay is that the length of the amplicon may affect the intensity of the amplification; thus, the present study designed primers for an amplicon less than 120 base pairs.

The World Health Organization and Pan American Health Organization published a recommendation for general procedures for inactivation of potentially infectious samples with Ebola virus and other highly pathogenic viral agents, indicating that using TRI reagent offers excellent performance and a safer environment for handling pathogens in a biosafety level 2 laboratory [10]. This is because TRI reagent has been shown to denature and destabilize the viral envelope and eliminate cellular nucleases while maintaining the structure of RNA for subsequent analyses.

The total cost for the SYBR Green assay was estimated to be $21, and the cost for the PCR was estimated to be $16. Although the PCR assay was more cost effective than the SYBR Green quantitative RT-PCR assay, PRC is more laborious and requires a longer time to complete (approximately 7 hours) than the SYBR Green assay, which requires approximately 4 hours. The time is longer for both techniques than that for using a commercially available detection kit because the two assays developed here are conducted manually. However, the cost for any commercial kit is considerably higher. Of the two manually conducted assay options we developed and validated for detecting SARS-CoV-2, we believe that the SYBR Green assay using RT-PCR is the more favorable choice owing to its potential power in the field of virology, the speed of conducting the assay, and real-time data analysis. The PCR assay was developed and validated as an alternative approach in the event that a laboratory does not have an RT-PCR instrument [11].

In conclusion, we developed and validated two low-cost assays for detecting SARS-CoV-2 that can be used by laboratories that cannot afford or cannot obtain commercial testing kits. Heating samples and using SYBR-Green or PCR assays might be not the ultimate solution for sensitive and accurate diagnoses; however, at a time when SARS-CoV-2 is globally pervasive and deadly and commercial kits are limited, these assays offer viable alternative testing approaches. In addition, these protocols provide virus testing platforms for future threats because the only change that would be required to follow the same protocols to test for a different virus would be the pathogen sequence needed to design specific primers. Furthermore, the protocols have been approved by the Saudi Food and Drug Administration as Emergency Use Authorization for use in a clinical setting.

## Data Availability

Any needed data will be available upon request

## Acknowledgments

We are grateful to the Department of Pathology and Laboratory Medicine chaired by Dr. Fouad Aldayel and to staff member Ms. Sarah Altamimi, who provided the deidentified patient samples. We thank Mr. Paul N. Muiya for synthesizing the PCR primers in less than 16 hours. We also thank Dr. Raed A. Abu Dawud from the Department of Comparative Medicine for his technical support. We extend our gratitude to the Sequencing Core Facility, Department of Genetics. The support of the King Faisal Specialist Hospital and Research Center (KFSHRC) administration is highly appreciated.

## Author Contributions

FA developed the study design, interpreted the results, and wrote the manuscript. AA developed the study design, interpreted the results, and reviewed the manuscript. DB worked on the primer synthesis. MB conducted the molecular assays. SA, MM, and RA provided the patient samples, ran the same samples at the diagnostic laboratory, and participated in the validation process and obtaining the Saudi FDA approval. DO completed the data analysis. All authors have read and approved the final draft of the article and have approved its submission for publication.

